# Country-specific optimization of testing rates and unlock measures can help to contain COVID19 infection

**DOI:** 10.1101/2020.05.20.20107169

**Authors:** Uddipan Sarma, Bhaswar Ghosh

**Affiliations:** Vantage Research, Sivasamy St, CIT Colony, Mylapore, Chennai, Tamil Nadu 600004. India; Center for Computational Natural Sciences, International Institute of Information Technology, Hyderabad 500032, India

## Abstract

In response to the COVID19 outbreak many countries have implemented lockdown to ensure social distancing. However, long lockdowns globally affected the livelihood of millions of people resulting in subsequent unlocks that started a second wave of infection in multiple countries. Unlocking of the economies critically imposes extra burden on testing and quarantine of the infected people to keep the reproduction number (*R*_0_) <1. This, as we demonstrate, requires optimizing a cost-benefit trade-off between testing rate and unlock extent. We delineate a strategy to optimize the trade-off by utilizing a data-trained epidemic model and coupling it with a stochastic agent based model to implement contact tracing. In a country specific manner, we quantitatively demonstrate how combination of unlock and testing can maintain *R*_0_ <1.

## Introduction

Declaration of the coronavirus pandemic by WHO severely overhauled global economic and social endeavors for more than 6 months now [1]. In response to initial upsurge of the infection spread [3,4], many European and east asian countries were able to contain the first wave successfully by imposing strong mitigation measures through nationwide lock-down coupled with rigorous testing and quarantine strategies. However, even with nationwide lock-down, many countries are struggling to contain growth of the infection[2]. The vaccine preparation is already on it’s way at a breakneck pace[5],but until then, isolating the infected population by aggressive testing and maintaining strong social distancing measures are adopted globally as the two most effective ways to deal with the current situation [6,7]. Presumably, despite benefits, long lockdowns are not sustainable approaches that severely disrupted the livelihood of millions, especially those of low and medium income people [8-11] calling for subsequent action plans to open up the economic activities. The unlocking measures subsequently led to an increase in infection rates and initiated the next wave of infection outbreak in multiple countries that were successful in their attempt to contain the first wave. For instance, second wave of infection is already being encountered in multiple regions of Europe and Australia. This inevitably calls for an immediate improvement in the strategies of testing and quarantine in order to maintain the basic economic activities while at the same time also ensuring a minimal spread of infection. However, the number of testing a country can conduct per day is limited by the resource capacity for testing and until now the maximum achieved capacity stands at around 5 daily tests/1000 globally [12] although countries have already taken initiatives to ramp up testing capacities [13-15]. Consequently, the allowed extent of unlock may also critically depend on the testing capacity. It is unknown how testing facilities and extent of allowed unlock for a country are related. This requires deciphering a country-specific cost-benefit trade-off that can aim at devising quantitative measures for optimally allowed extent of the unock in view of the maximum resource capacity for testing. In this global emergency scenario, and in the absence of vaccines, quantitative understanding of the relation between testing rate and an respective optimal unlock extent can potentially contain the infection spread.

Here, we investigated the trade-off using a dynamic epidemic model [16-19] for the spread of coronavirus. By quantitatively calibrating the time series data [1] for confirmed, recovered and dead population for 50 different countries with various stages of infection, we made an estimate of parameters like incubation time, transmission rate, quarantine, recovery and death rates. We introduced lock-down in our model to observe the effect of social distancing and also estimated the effectiveness of implementation of lock down in individual countries. Using the fitted parameters from the dynamic epidemic model, an analytical calculation was next performed to determine the reproduction ratio (*R*_0_) of the model and on the basis of that calculation we develop a model to implement the trade-off between different extent of partial unlock and the quarantine rate allowing us to find optimum extent of allowed partial unlock which would maintain low *R*_0_<1, given a maximum capacity of quarantine rate. One way to implement partial unlock is to have periodic unlock with a certain frequency [20] and here we connect the extent of unlock with the frequency of the unlock time over a period of two weeks. However, the ordinary differential equation (ODE) based model can not explicitly capture the relation between infection rate (*R*_0_) and testing rate. To introduce testing, we developed an agent based stochastic version of the epidemic model explicitly incorporating testing through contact tracing of individual infected agents at every time step. The calculations in the agent-based model are made country specific by selecting the fitted parameter values from the ODE based model specific to a country. Our study thus provides an optimum strategy to connect the testing/1000 population necessary to allow a particular extent of unlock in a given country which individual countries can use for devising unlocking approaches based on their testing facilities.

## Results

### Doubling rate of the infection is determined by the testing rate

To explore the dynamics of the COVID19 infection spread we took the daily confirmed infection time course data from WHO and clustered the region-wise data according to their dynamic pattern. A hierarchical clustering algorithm (hierarchical clustering from pheatmap package in R) is used to analyze the dynamics of around 100 countries selected (countries with at least 1000 infections per day in their maximum infection spread phase). The provinces in China clearly are clustered together since infection spread happened at the earliest times (Figure 1A). Then the infection spreads to different parts of Western Europe followed by East asian and south american countries which can also be seen as close proximity in the clustering analysis (Figure 1A, color bar represents normalized daily confirmed cases). Next, we calculated the doubling rate from the time series of the countries. The doubling rate is defined as the inverse of the doubling time, i.e., how much time does the population take to double the number of infections (Figure 1B). The doubling rate time courses for the different countries are then aligned at the maximum doubling rate and we calculate linear correlation at different timepoints with the number of tests/1000 population for individual countries. The same calculation is performed for the daily confirmed cases. The normalized daily cases shows a positive correlation with the test rate around a time point of 16 days (Figure 1C, S1B, 0 corresponds to time point corresponding to maximum doubling rate), suggesting, countries with higher test rates tend to have higher confirmed cases. The doubling rate after around 26 days displays a negative correlation with test rate (Figure 1D, S1A lower panel) indicating countries with higher testing rate are able to reduce the rate of infection spread faster. However, it is noticeable that, during the start of the infection doubling rate is usually high for higher testing rate as higher testig rate (Figure S1A top panel) allows more precise measurement of the infection spread which can later facilitate containment of the infection through preventive measures such as quarantine. However, notably the testing strategies in most of these countries are implemented under lockdown conditions. In order to project required enhancement testing w.r.t to different degrees of removal of lockdown we take recourse to a quantitative epidemic model and subsequently an agent based model.

**Figure 1:**
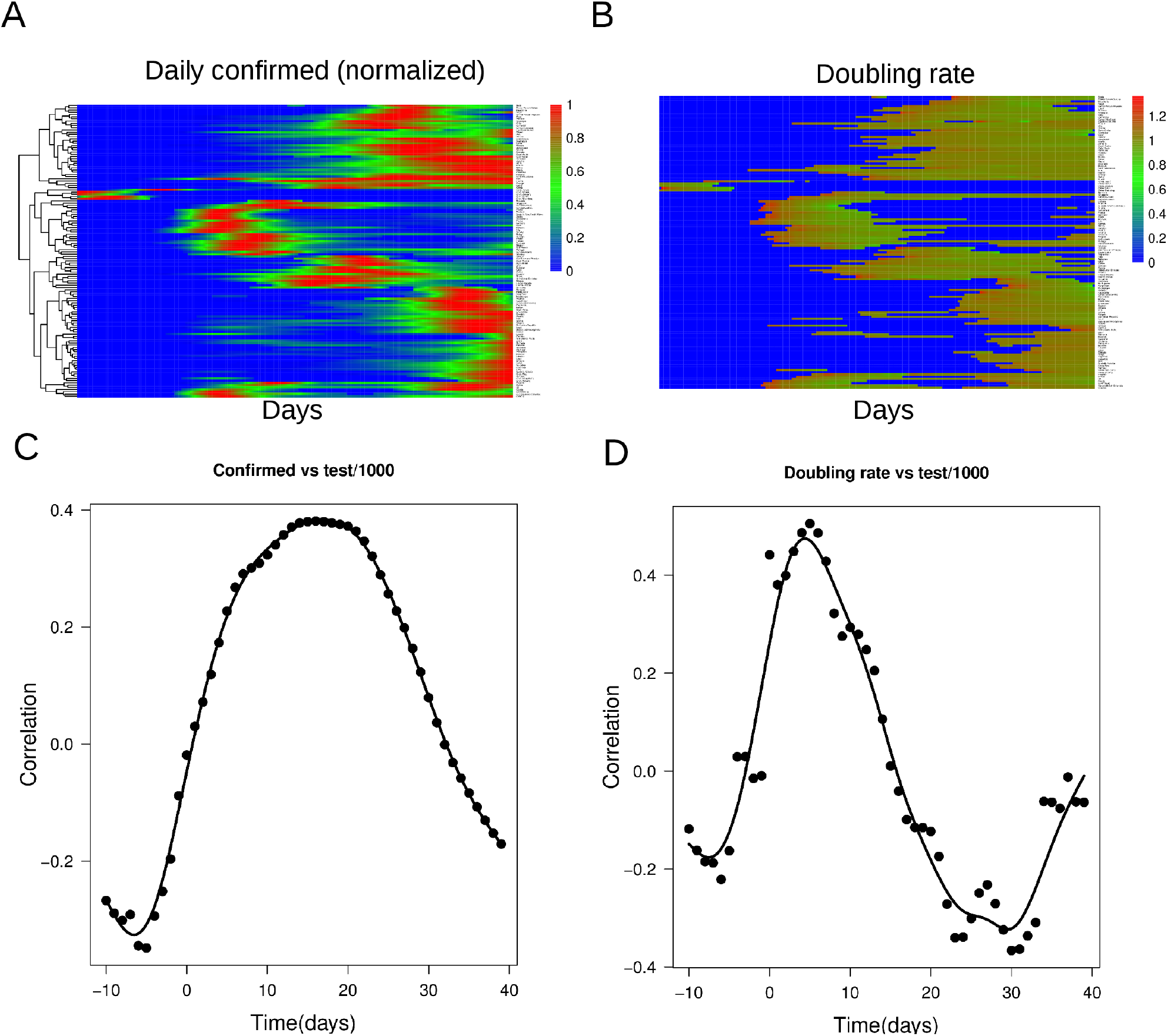
The Covid-19 outbreak in different countries and its relation with the testing rate. (A) The heat map displays the clustered dynamics for around 100 different countries for the daily cases normalized to the maximum for each country (Days correspond to 22^nd^ January, 2020 and to 15^th^ August, 2020). The dendogram is based on hierarchical clustering of the time traces (B) The heat map represents the doubling rate as a function of time clustered according to the dynamics of the time traces as in (A). The results are shown in exponential scale. (C) The correlation of test rate with the daily confirmed cases at different time points along the dynamics are shown. The time traces are aligned w.r.t the maximum doubling rates. 0 on the x-axis corresponds to the maximum doubling rate for all the countries. (D) The correlation of test rate with the doubling rate at different time points along the dynamics aligned as described in (C).

### A compartmental epidemic model is calibrated to obtain country specific infection parameters

From the group of 100 countries, we selected 50 representative countries, comprising a combination infection stages and infected population size, and fitted their confirmed (Co), recovered(Re) and dead(De) population trajectories (Methods for details). The model contained five compartments namely susceptible (S), exposed (E), Infected (I), Quarantined (Q) and removed (R), where R contains both recovered and dead population [21]. Figure 2A shows the structure of the SEIQR model. Here a susceptible person can be exposed to the infection through transmission from an infected person. After exposure, the symptoms are exhibited within an incubation time and the infected individual either recovers or dies after a time, represented by a recovery or death rate. Dynamics of the system are captured by a set of 6 coupled ordinary differential equations (details in methods). The calibration data for each country comprises the time courses of the number of Co/Re/De and through model fitting we estimated the parameters that best explains the Co/Re/De trajectories simultaneously of each country. Model fitting also includes a lockdown function [22]. The lock down is introduced in the model through a reduction in the transmission rate that follows an inverse sigmoid function. The process of lockdown is controlled by three variables-time of lockdown (start time of lockdown implementation), strength of lockdown (the extent of lockdown in a country, 0.1 would mean 90% lockdown) and the effectiveness of lockdown (how fast the maximum lockdown is achieved from the lockdown starting time point); during model calibration these parameters were estimated in a country specific manner. Figure 2B shows the model fits the confirmed, recovered and dead populations of 4 representative countries. Similarly the Co fits to all the other countries are shown in supplementary Figure S2, respectively. Here (Figure 2B) we show the cumulative trajectories for Co/Re/De simultaneously. The confirmed trajectory by definition comprises [quarantined + recovered + dead] population and the infection (I) compartment represents the unidentified/unknown infection cases present in the population. The assumption is rooted in the fact that once an infected person is tested positive he/she will be quarantined, thus, only the infected but untested subjects (I) can further infect the susceptible. It is notable that the average fitted incubation time comes out to be around 8.1 days which matches with the observed value [23].

**Figure 2:**
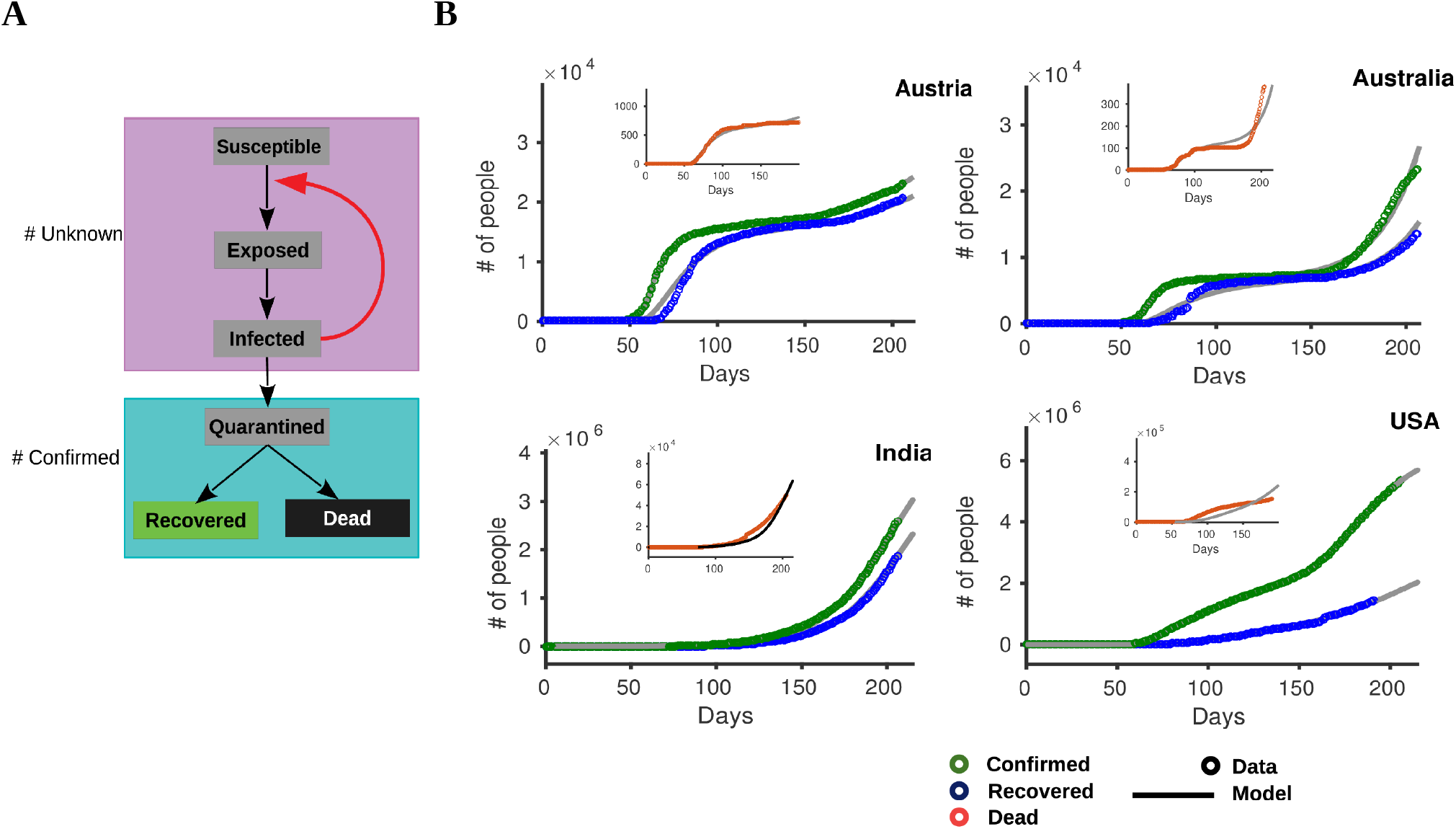
The description of the SEIQR model and calibration for different countries. (A) The model utilized to fit the cumulative confirmed, recovered and dead cases comprises susceptible, exposed, infected, quarantined, recovered and dead compartments. The lockdown is implemented through a sigmoid function as indicated. The quarantined, recovered and dead cases together comprise the confirmed cases. (B) The SEIQR models fit the data for 4 representative countries as indicated for the cumulative confirmed, recovery and death cases. The number of days in X axis corresponds to the time course data available in JHU CSSE [39] where 0 coroposponts to 22^nd^ January, 2020 and the end time point corresponds to 15^th^ August, 2020.

Next, we obtain *R*_0_ value for all the countries by capturing the dynamics of the lockdown function. The calculation shows a wide variety of responses among countries with respect to the effect of lockdown. Some countries are able to reduce the *R*_0_drastically after lockdown whereas in some countries impact of lockdown is not very high (Figure 3A). The test rate displays a negative correlation with *R*_0_after lockdown (Figure 3B), but correlation is not significantly high. The infection rate *R*_0_is dependent on many other factors including demographics, medical facility and distribution of virus strains in the population which may influence the impact of testing rate. In order to check the influence of these factors on *R*_0_, we collected several publicly available datasets of demographics, medical facility and genome sequences. The *R*_0_ values show significant correlation with some factors including median age, life expectancy, population size, doctors/10000 (Figure 3C,S3A). Some significant correlations with frequency of the clades in the population were also observed. Specially the L and GR strains shows significant influence on *R*_0_(Figure 3D, S3B). We now linearly adjusted the *R*_0_ values (Methods) using the most significant factors derived from the correlation analysis, namely median age, population size, doctors/10000 and frequency of the L clade. We found, the adjusted *R*_0_values exhibit much higher correlation with the test rate (Figure 3E) corroborating our emphasis on test rate and quarantine in containing the infection. We would like to mention that for the correlation analysis, we choose countries only with at least 10 complete genome sequences till the end of June which selects 39 countries out of the 50 countries. To this end, with the fitted parameter values for all the countries the effect of the quarantine and test rate on the infection spread after removal of lockdown can be explored. A specifically selected 5 representative countries with high transmission rates (USA, India), low transmission rates (Australia, Austria) and high transmission rate and high quarantine rate (France) are next studied to capture a generalized nature of different infection spread scenarios (Figure 3F).

**Figure 3:**
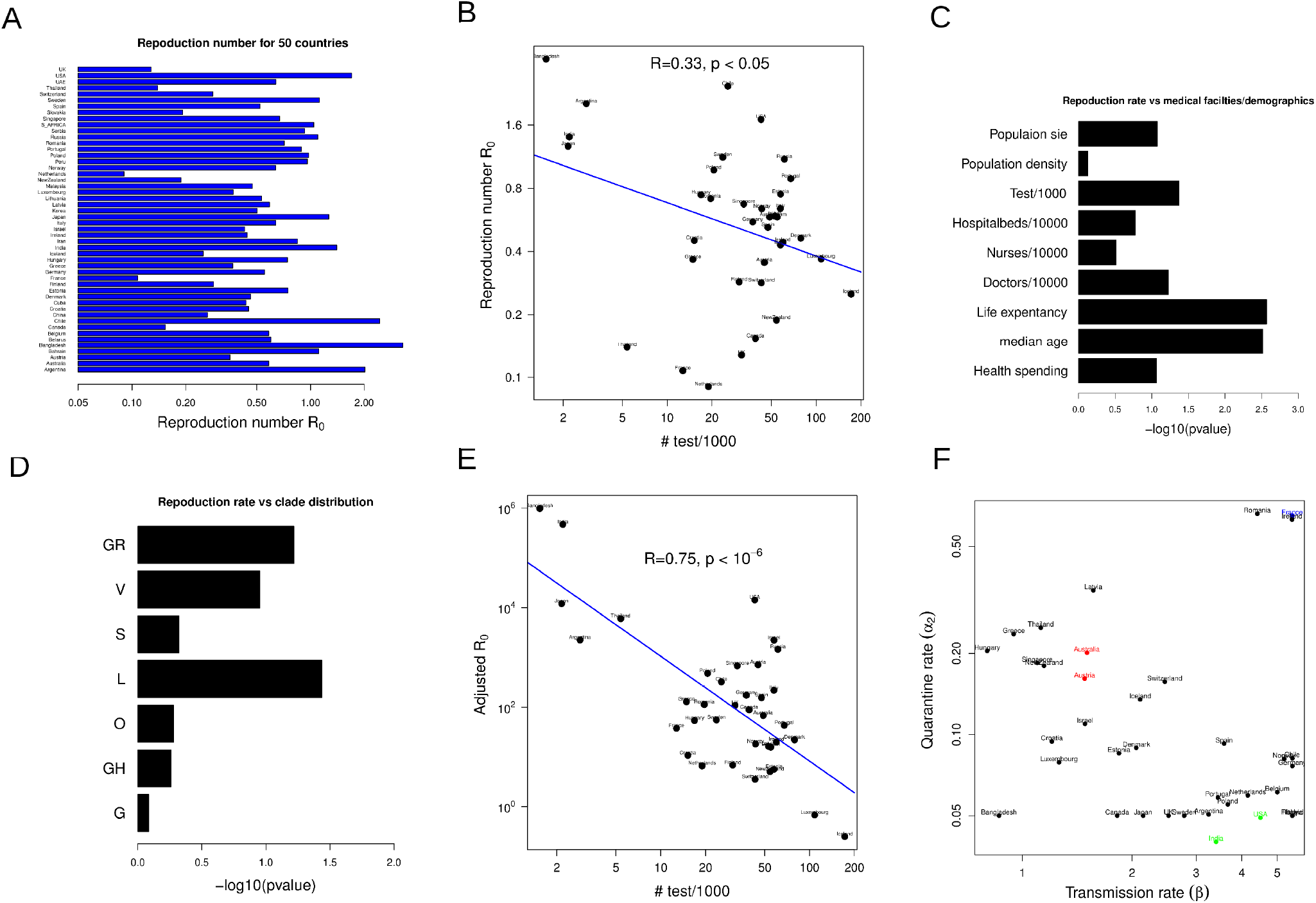
The reproduction number (*R*_0_) displays correlation with the testing rate. (A) The barplot shows *R*_0_ for the fitted 50 countries after lockdown. (B) The scatter plot describes correlation between the test rate and *R*_0_after lockdown for the 39 countries with at least 10 complete genome sequences as of end of June described in the main text. (C) The statistical significances of the dependence of *R*_0_ on different demographic and medical facilities factors are shown based on linear regression based on the same 39 countries (D) The same statistical significance of dependence of *R*_0_ on the frequencies of the clades (G,GR,GH,L,O,S,V) are shown. (E) The scatter plot indicates the correlation between the adjusted *R*_0_ values and the test rates. (F) The values of the fitted transmission rates and quarantine rates for all the 50 countries are plotted and the representative countries for consideration of the agent based model simulation are shown in colors red, green and blue.

### Optimum quarantine rate change, uniquely for each country, can effectively reduce the infection post lockdown

Typically, when lock down is removed, the transmission rate (β) would increase substantially. For the SEIQR model we considered here, the *R*_0_ value is given by 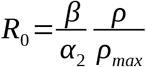. where *ρ* captures the lockdown. Thus, from this expression we can calculate the extent by which the quarantine rate (*α*_2_) must be increased in order to compensate for the effect of partial or full unlock. The fold change in *α*_2_ to keep the *R*_0_ value close to one is given by

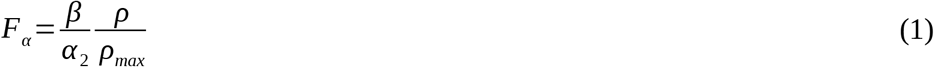

Here, *ρ* = 1 corresponds to fully lockdown case and gradual increase in value of *ρ* depicts partial unlock finally moving towards full unlock at the fitted value of *ρ_max_*. The equation clearly shows that the required fold change to compensate for the effect of lockdown removal would reduce as the extent of unlock reduces(Figure 4A). In fact, according to the fitted values, the transmission rate has higher value for the USA compared to Austria, Australia or India leading to a higher required quarantine rate for the same unlock (Figure 4A). However, for France in spite of very high value of *β*, the requirement for increased fold change in quarantine is low due to it’s high quarantine rate (Figure 4A). Thus, the removal of lockdown imposes an extra cost to the country in the form of a requirement of increased quarantine rate which can only be achieved by rigorous contact tracing and testing. This scenario can be thought of as a cost-benefit tradeoff where on one hand removal of lockdown benefits economic activities but at the same time more testing needs to be conducted to circumvent the enhanced infection spreading rate. If we assume that the maximum quarantine capacity of fold change in *α*_2_ is *F_c_* and investment cost in testing is *λ* per testing, the optimal fold change in *α*_2_ would be (methods)

**Figure 4:**
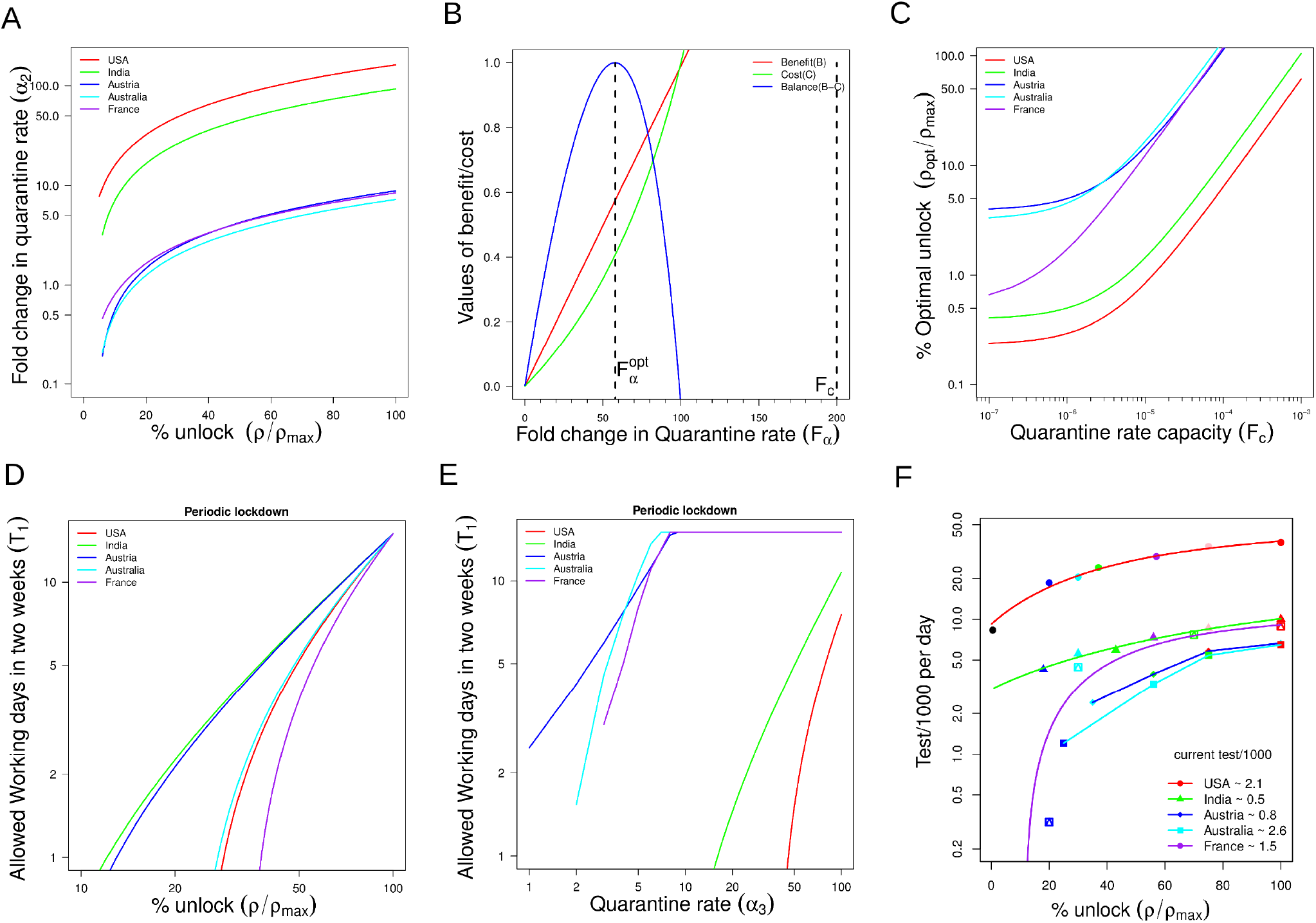
The optimization of trade-off between testing rate and extent of unlock. (A) The relationship between the extent of unlock and the corresponding quarantine rate required to keep *R*_0_ values equal to one for five countries as indicated. (B) The cost and benefit curves as a function of quarantine fold change for the countries. The cost benefit is optimized at a value 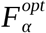 for a maximum capacity *F_c_* of the fold change in quarantine rate. (C) The optimum extent of unlock calculated from 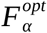 as a function of the maximum 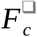. (D) The implementation of the extent of unlock as a function of the periodic unlock for the different countries are indicated. Number of working days represents the working day cycle in 15 days time. (E) The quarantine required to be increased by testing to keep the 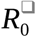 value below one and the corresponding the working days allowed. (F) The allowed extent of unlock and the corresponding test/100 required to keep the 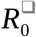 value less than one calculated from the agent based simulation for the different counties with indicated current test rate.

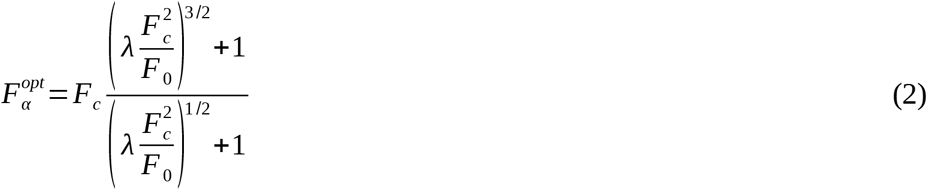

Figure 4B shows an illustration of the optimality in quarantine rate given a maximum capacity of quarantine rate. Finally from equation (1) the optimal lockdown removal (*ρ_opt_*) is given by

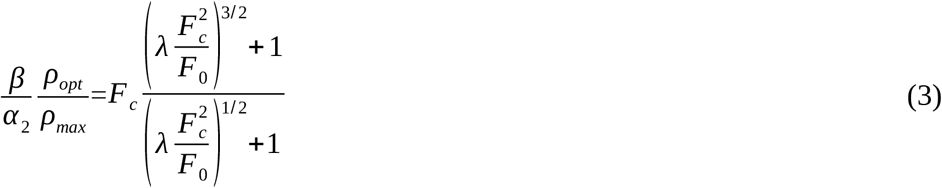

This equation reveals that as the quarantine capacity *F_c_* is low, the *ρ_opt_*would be high (Figure 4C) which illustrates the fact that as the quarantine capacity is low, the extent of lockdown should optimally remain high. Even though, higher *F_α_* value is possible, total benefit would be low given the cost. Since the USA has a higher value of *β*, given a capacity of quarantine rate, the optimal unlock is low for the USA (Figure 4C) compared to Austria, Australia which have much lower values of *β*_❑_. Similarly, it also shows that the optimal extent of lockdown would increase if the cost of testing increases. However, we would like to mention that the extent of lockdown can be effectively implemented through a periodic lockdown with a period of *T*_1_period followed by *T*_2_ of lockdown period [20]. In this case, one can show that the allowed unlock period, the average *R*_0_ would be given by (Methods)

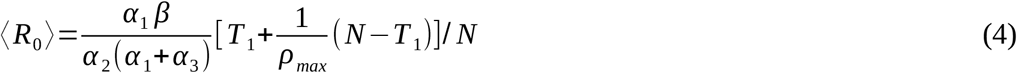

The frequency of the unlock can be connected to the extent of unlock through the equation below

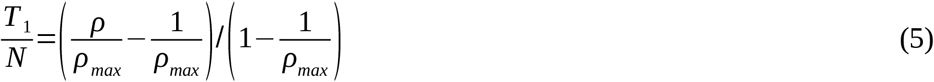

This shows the frequency of unlock required in a time N to implement a particular extent of overall unlock 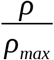 (Figure 4D). The corresponding unlock period to keep the average value at 1 is given by

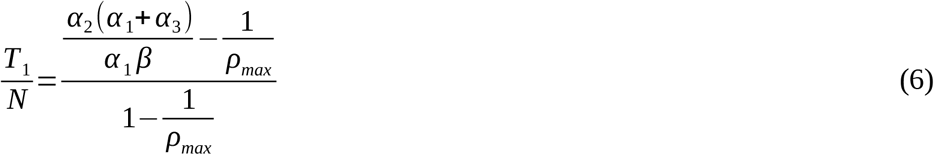

which represents allowed unlock number of days over a period time N to keep the average *R*_0_ at equal to 1. This shows that as the effect of lockdown *ρ_max_* is high the allowed unlock period would also be high. The allowed period can further be stretched by increasing the quarantine rate (Figure 4E) involved with testing and quarantining the exposed subjects at a rate *α*_3_ where as small value of *α*_3_ corresponds to the case when testing rate is low which is used in fitting the data.

However, this ODE based model is not sufficient to quantify the number of testing required to achieve a certain quarantine rate *α*_3_. In order to get a grasp of the number of testing, we have to introduce testing through contact tracing in the model which is not possible to explicitly implement in an ODE based model. To achieve this, we constructed an agent based stochastic version of SEIQR model where the parameter values are selected from the already fitted values from the ODE model (Methods). The infection spreads outward starting with one infected agent at the centre. The selected model for the parameter values of the USA demonstrates that as the testing rate increases the maximum confirmed cases also increases (Figure S4A) as expected from Figure 1C, but the infection is contained much earlier leading to a reduction in the reproduction number for different partial unlock cases (Figure S4B). The results are reproduced for other countries as well (Figure S4C-F). From these figures, we used an extrapolation method based on cubic spline fitting to determine the minimum test rate required to keep the *R*_0_ value less than one for each partial unlock case as indicated in the figure. This calculation results in an expected monotonic increase in the required testing rate as the extent of unlock gradually progresses from full lockdown towards full unlock cases (Figure 4F) although the exact values of required testing rates vary among countries for different partial unlock depending on the transmission and initial quarantine rates (Figure 4F). It is noticeable that due to low transmission rate without lockdown countries like Austria, Australia is able to unlock by 40% with a very small ramping up their current testing rate. Whereas for USA, India the current testing rates need to be ramped up at least by 10 times to achieve an unlock of 40%. A 40% unlock corresponds to a periodic unlock of 5 days of working days followed by 10 days of lockdown period according to Figure 4E. So, to summarise, the USA needs to increase its testing rate by at least 10 times in order to proceed to 5 working days in 15 days. On the other hand, Austria/Australia can open their economy by 5 working days with a less than 2 fold change in their current testing rates. We argue that the burden on the testing rate capacity can be essentially reduced by keeping the transmission rate low through other measures of social distancing, wearing face masks etc.. Thus, in conclusion, in order to leverage the benefit of augmented testing capacity, the population must practice other social distancing measures which would keep the natural transmission of the infection low.

## Discussion

In this paper, using an epidemic model, we quantitatively explored the COVID19 outbreak and its relation with quarantine measures in different countries. The reproduction numbers of this pandemic are found to be comparable to the SARS-cov values [24,25] and much higher than the MERS infection[26,27]. We employed an epidemic model and fitted the confirmed, recovered and dead population trajectories from 50 countries where the countries are a combination of different stages of infections. Some countries are yet to control the first wave of infection whereas in some other countries the second wave of infection has already begun. We built an agent based stochastic model [28] to implement contact tracing where we utilized the parameters quantitatively derived from calibration of Co/Re/De dynamics, in a country specific manner. The primary goal of utilizing the two distinct modeling approaches is to connect the testing rate to the unlock process, where, and either of the modeling framework cannot connect both as ODE based epidemic models alone cannot implement the contact tracing. Indeed, previous studies discuss the limitations of ODE based models and agent based models [30] but none of the studies connected both the approaches in the context of COVID19 epidemics. Here, by connecting the ODE model with the agent based model, we derive country specific optimal testing-rate through contact tracing that is required to allow a certain degree of unlocking to open up the economy. The model can also potentially demonstrate the advantage of contact tracing over random sampling (data not shown). Lockdown is a preparatory measure for the health care system to reorganize itself to deal with the situation since long term lock down would be detrimental to the economy of any country[29].

Typically the actual infected number of people is expected to be higher than the sampled one’s and limitations like that pose challenges for devising accurate mathematical models [30]. Actual susceptible population size is thus mostly unknown and it could often be different from the tested population size, in either direction. As a result, the number of infected people from the data only captures the infected people out of the tested sample. This is one of the reasons why increased testing rate is so important in capturing the real magnitude of the infection (and not only the dynamics) that would also lead to devising more accurate epidemic models with better predictive capacities as underpinning the size of true susceptible population in an infected country is key to improving model forecasts on the disease spread. In addition, because of the asymptomatic nature of the infection [31], the need of quarantining infectious people through testing is even more important to successfully contain the infection spread. However, it can be also be noted that, apart from the testing rate, other factors like immunization [32], age [33], sex [34] medical facility and specific virus strains [35] may play important roles both in transmission and fatality.

In countries like India with high population density and size and with a relatively inadequate health infrastructure, an early implementation of lockdown post infection was critical in adequate containment of the infection spread, whereas, the delay in lockdown such as in Italy or Spain had a much more catastrophic impact despite adequate health infrastructure; highlighting the importance of early measures during such outbreaks. This is further highlighted at present where unlock measures chosen by countries like India and USA immediately witnessed dramatic increase in infection spread. However, reopening the economy is also an impending necessity in all countries under lock down. Thus, to minimize the health hazards of social proximity while being able to continue economic activities, will require data-driven planning and implementation. We propose strategies where rigorous quarantining of the infected subjects is argued as the only effective measure to successfully deal with infection spread post-lockdown and pre-vaccination phase. As a policy measure, our model suggests that quarantine and testing should be increased substantially after lifting the lockdown, in order to contain the infection in the coming months. We demonstrate, the effective increase in quarantine measures is country specific, depending on the transmission or quarantine rates. Countries have taken initiatives to accentuate the testing capacities either through developing more testing facilities [13-15] or through designing efficient pool testing algorithms [36,37]. We believe our study will ease planning of such testing processes where, for instance, if a goal of 30% unlock is set by a given country(or a province in a country), then the respective increase in testing rates to achieve such unlock goals can be predefined as a minimal testing rate target. Although periodic lockdown [20] is an efficient way to implement partial lockdown, we also argued that this would only work if the *R*_0_value during the lockdown period is kept much lower than 1, which requires testing and quarantining, as in many developing as well as developed countries it is almost impossible to limit the mobility of low income people even during lockdown [10]. A small unidentified fraction of the infected population during the lockdown removal can potentially remain unidentified due to the long incubation period characteristic to COVID19 infection. Indeed the Spanish flu pandemic in 1918 came back again in a few weeks after the first wave was apparently contained, with a more catastrophic impact[38]. The cost-benefit approach we derived shows a data-driven strategy to achieve *R*_0_ <1 at all times and thereby bypass such major uncontrolled future outbreaks, implementation of which can be tailored in a country specific manner.

## Data Availability

Details of the model simulation and fitting are available upon reasonable request.

## Author contribution

US and BG conceived the study. US built the epidemic model, developed fitting and prediction strategies and optimized the parameters. BG developed the agent based model, performed the stochastic simulations, analytical calculations, genome sequence analysis and regression analysis. US and BG analyzed the results and wrote the manuscript.

Conflict of interest: The authors declare no conflict of interest.

## Supplementary Figure legends

**Figure S1:**
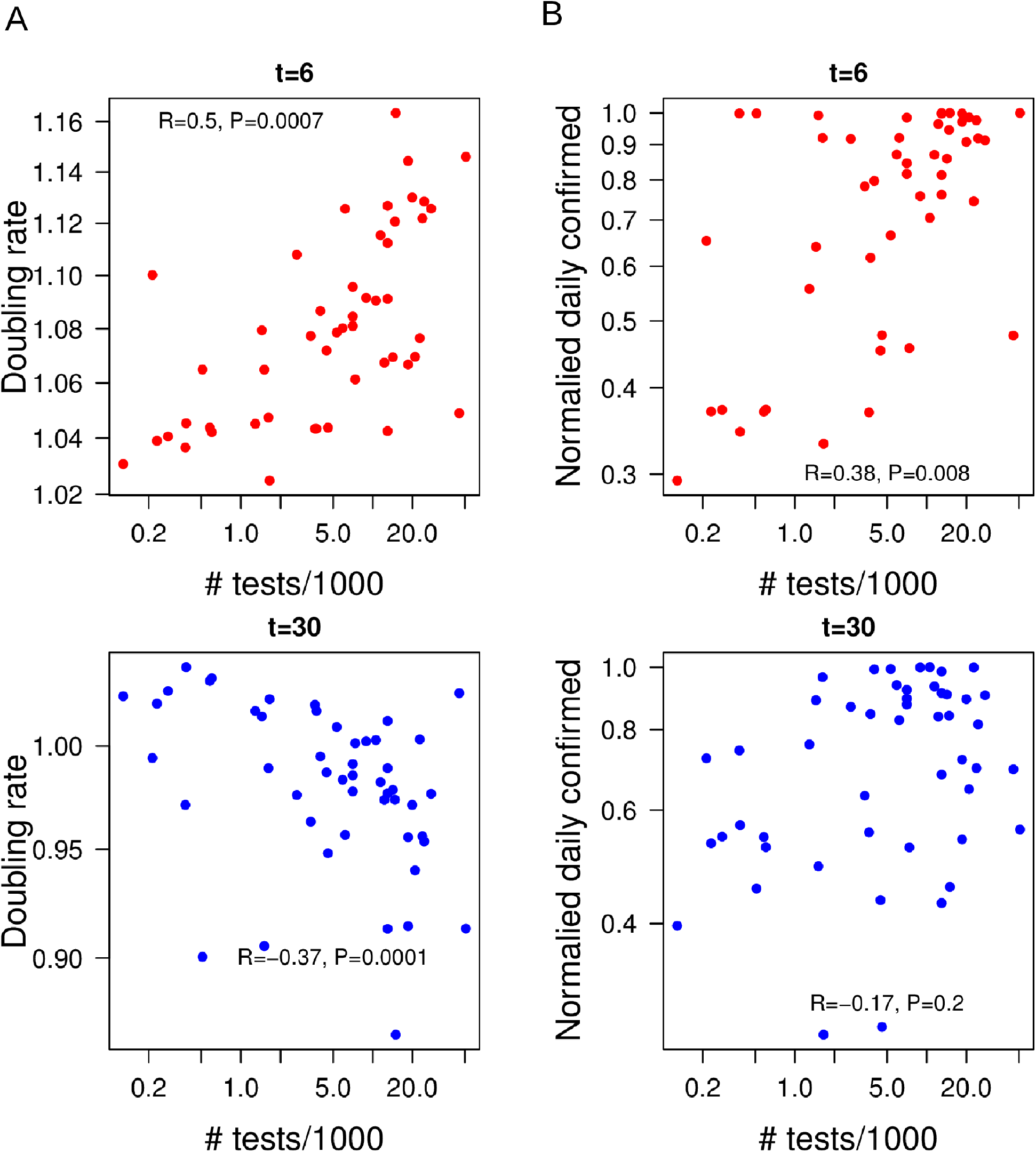
The scatter plot at two different time points between test rate and daily confirmed cases ase well as test rate and doubling as an example for generating the Figure 1C and 1D.

**Figure S2:**
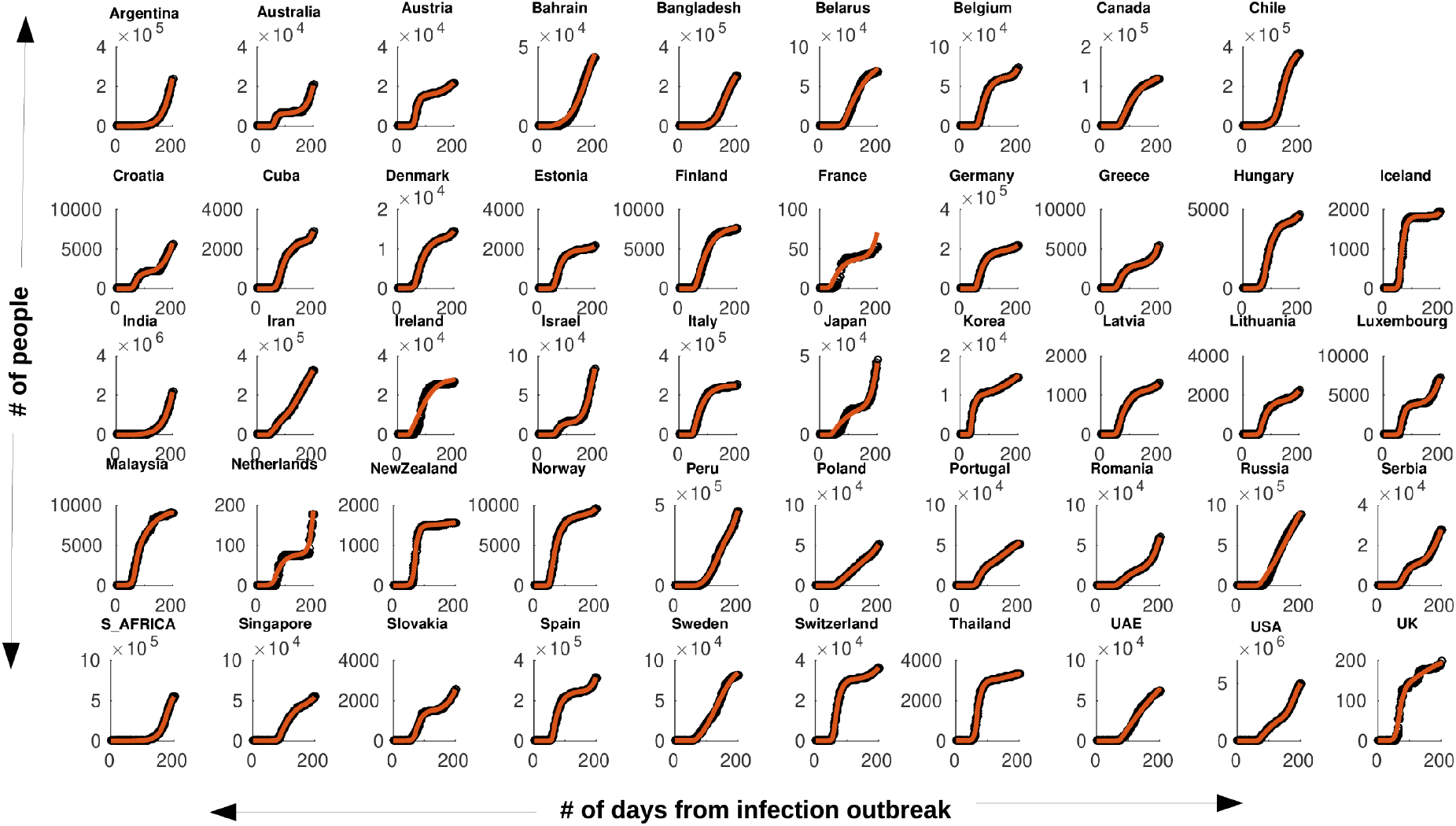
The SEIQR models fit the data for 50 countries, as indicated, for the cumulative confirmed cases. The figures indicate the cumulative data and the corresponding fit based on the SEIQR model.The number of days in X axis corresponds to the time course data available in JHU CSSE [40] where 0 coroposponts to 22^nd^ January, 2020 and the end time point corresponds to 15^th^ August, 2020.

**Figure S3:**
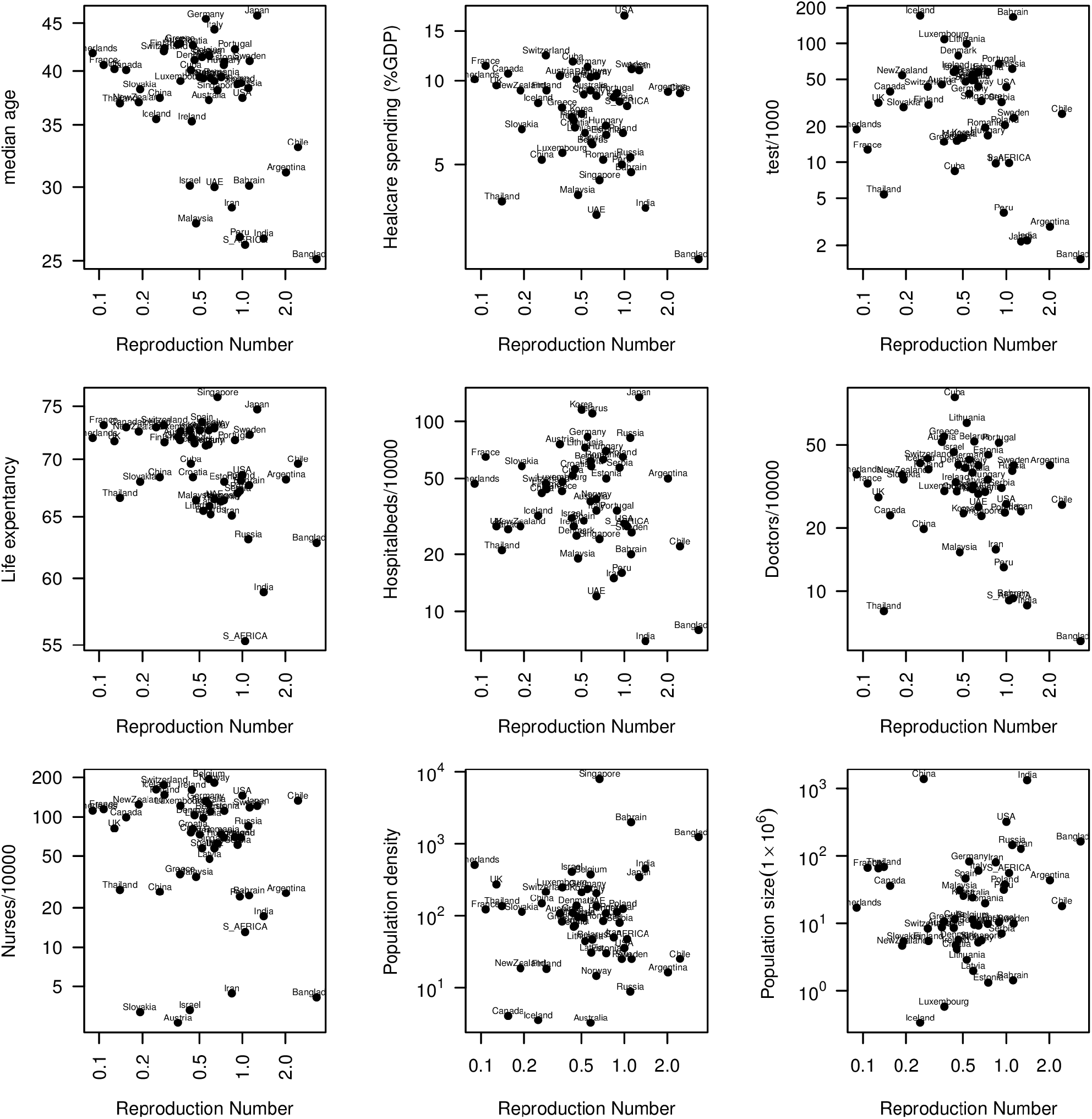
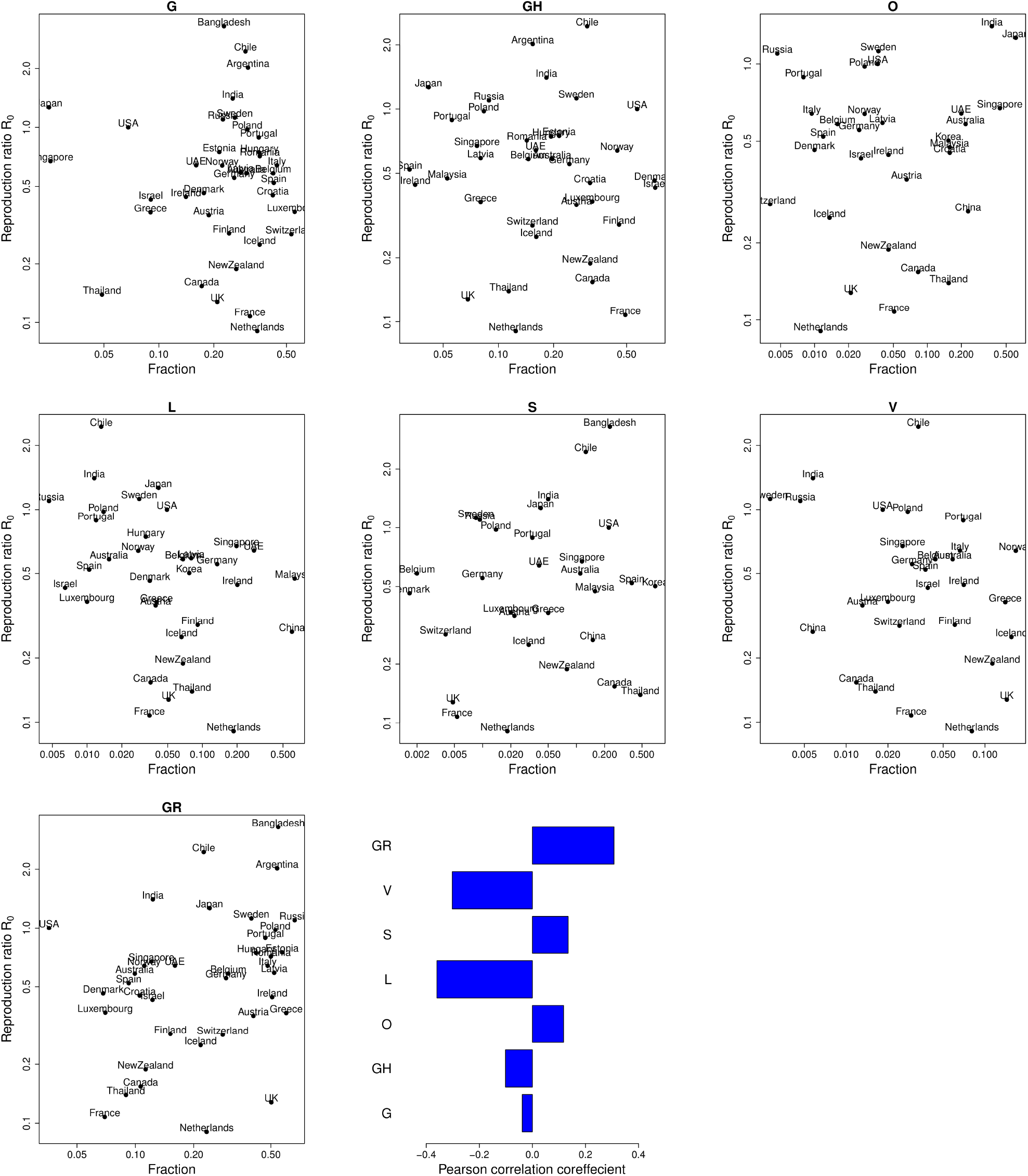
(A)The scatter plot showing the relationship between the *R*_0_ and different demographic and medical facility factors as indicated in the figures. (B) The scatter plot shows the relationship between the frequencies of seven clades in different countries and corresponding *R*_0_ values.

**Figure S4:**
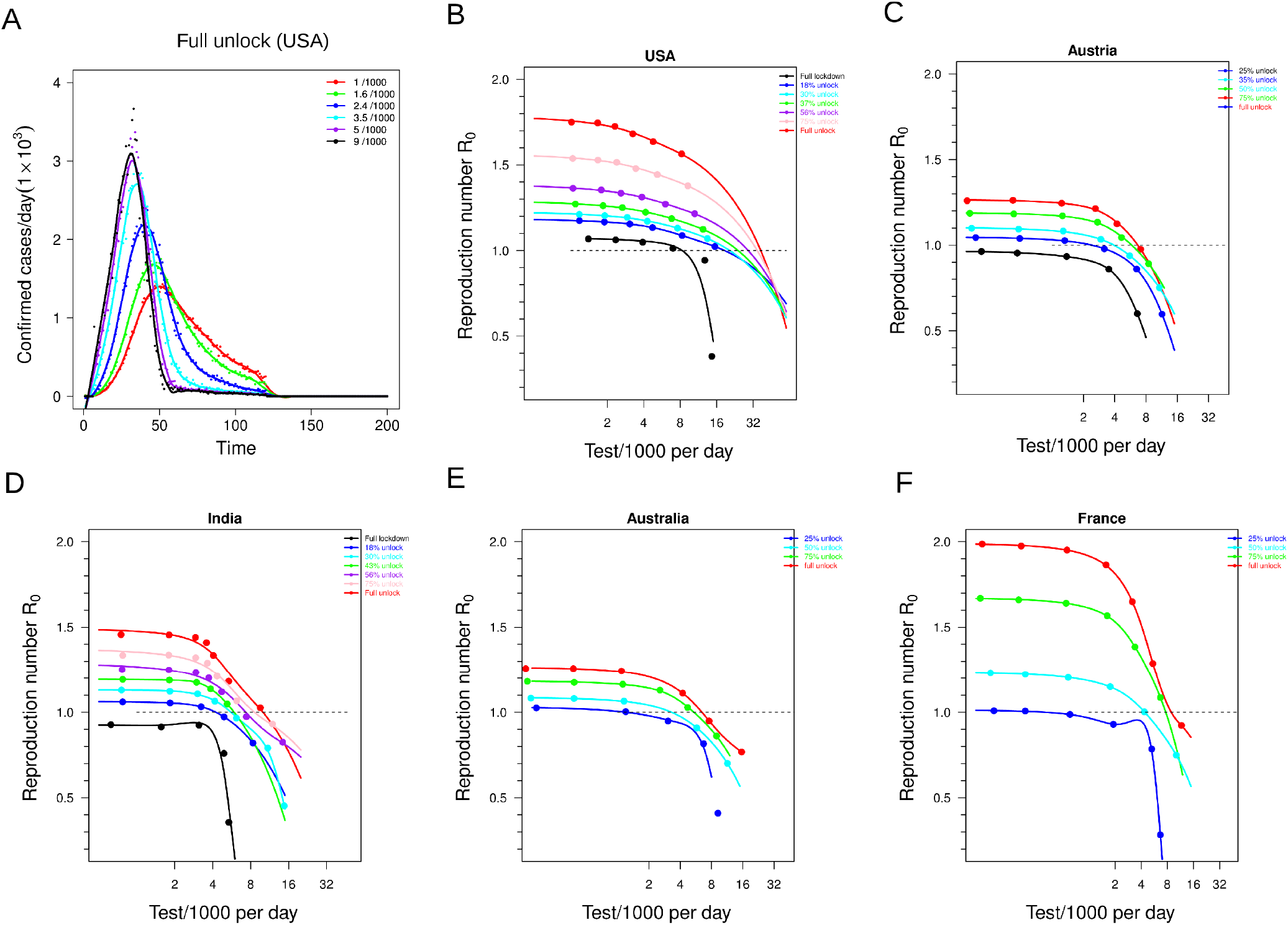
(A) The time traces of the confirmed daily cases of agent based simulation based on parameter values with full unlock for the USA at different daily tests/1000 as indicated. (B-F) The *R*_0_ values based on the agent based model with parameter values of the five different countries as indicated at different test/1000 rates. The dashed horizontal line exhibits the *R*_0_=1 line, intersection of the line with the curves quantifies the test/1000 required to maintain *R*_0_=1.

## Supplementary Information

### Materials and Methods

#### Calculation of doubling rate

The doubling rate is calculated at each time point over the trajectory by taking the daily cases at that time and next day. For an exponential growth of the infection, *N*(*t*)=*N*(*t* +1)exp(*λ*) where N(t) depicts the number of infected people at time t

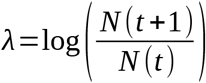

**The SEIQR model:** The model comprises of susceptible(S), Exposed(E), Infected(I), Quarantined(Q), Removed(R, contains two compartments ‘recovered’ and ‘dead’)

The equations are

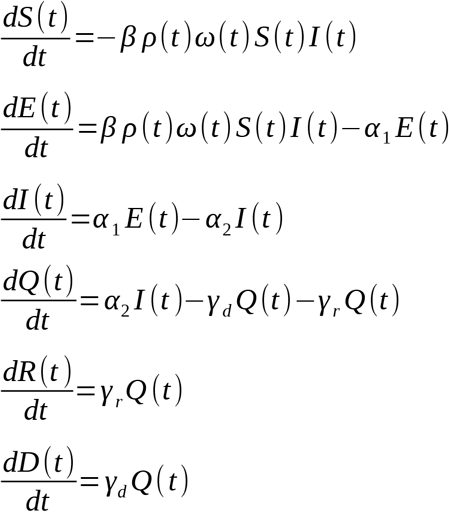

Where S(t), E(t), I(t), Q(t), R(t) and D(t) are the susceptible, exposed, infected, quarantine, recovered and dead population at time t respectively. ρ(t) is the lockdown function and *ω*(t) is the time when infection starts in a given country after the first day of detection of the infection date is reported in WHO website (22nd Jan, 2020).

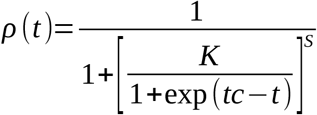

where, K=Effect_lockdown_Country, S=*strength_¿_*

and tc=*Time_¿_*

ρ(t) varies between 0 and 1(no lockdown) where 0 (full lockdown). *ω*(t) ensures that the model for a specific country is switched on when the infection begins in that country, hence if the detected case in a country is 40 days after Jan 22nd, the model for that country is switched on 35 days post Jan 22nd (assuming mean incubation time of 5 days), during the calibration.

The lockdown is opened by modifying the ρ(t) function such that ρ(t) returns to 1 from its lockdown status to no-lockdown(1) status in a designated time.

#### Model calibration

Model calibration involves minimizing an objective function that gives best fit parameter sets for confirmed, recovered and dead populations for a given country simultaneously. We fitted the time series provided by JHU CSSE at github [1] to the SEIQR model developed in the study and minimized the objective function using the lsqnonlin function of MATLAB which minimizes differences in the sum of squares between model and data. To fit the observed cumulative confirmed trajectory for a given country, [quarantined + recovered + dead] from the model is fitted against the confirmed data. The objective function is thus minimized to achieve the best fits for Co/Re/De simultaneously for a given country. This was repeated for all the 50 countries individually.

#### Analysis of the virus genome sequences for clade distribution

We first collected the individual sequenced genome from individual countries from [40] and their corresponding clades as reported in [2]. Each genome in a particular country belongs to one of the seven clades (G,GH,GR,L,S,V.O) as described in [40]. From the data, we determine the frequency of each clade in individual countries by finding the number of sequences belonging to that clade divided by the total number of sequences. To calculate the correlation, out of the analysed 50 countries, we only considered countries having at least 10 sequences in the database.

#### Adjusted *R*_0_values

The *R*_0_ values obtained by fitting are adjusted with respect to the significant parameters from the correlation analysis described in Figure 3C,D and S3A,B. The adjustment factor is characterised as

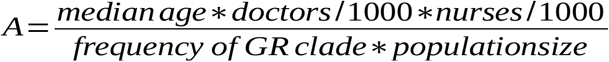

The factors which are positively correlated with *R*_0_ (population size, frequency of GR clade) are placed in the denominator and factors having negative correlation values are placed in the numerator. The adjusted *R*_0_ values are quantified as

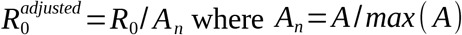

#### A model for optimizing the quarantine rate

The basic reproduction ratio for the SEIQR model discussed above is given by

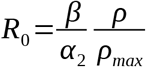 where 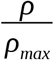 represents the extent of unlock. So this equation can be transformed as

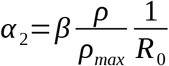

The value of quarantine rate *α*_2_ *‘* to keep *R*_0_ equal to one would be

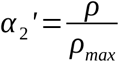

Thus,

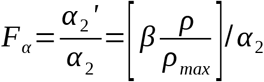

This equation shows that the required fold change to the keep *R*_0_ value lower than one is higher as the extent of lockdown is more. Thus, we argue that the higher *F_α_* has a benefit of unlocking the economy but it comes with a cost in spending financial as well as medical resources in testing. The benefit can be simply written as 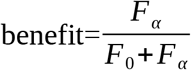 takes into account the fact that above certain high quarantine rate *F*_0_ the benefit would saturate. The cost, on the other hand, would also increase with higher quarantine rate as 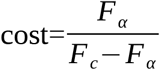 where *F_c_* depicts the maximum capacity of the quarantine rate possible for a particular country on the basis of it’s financial and medical resource limitations. The overall balance between the cost and benefit can now be written as 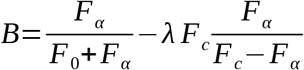 where *λ* represents the resource utilization price per unit of maximum testing rate.

At the optimal *F_α_* value, the derivative of the balance equation must be zero. Hence,

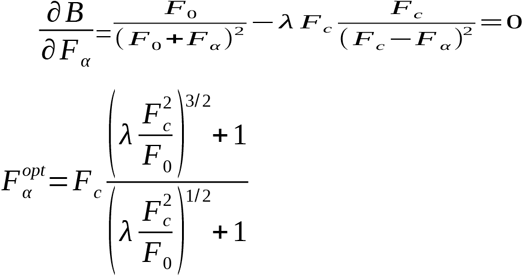

Thus the optimal unlock would be

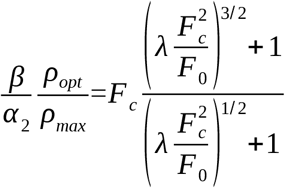

#### Agent based stochastic SEIQR model

We utilized the SEIQR model as described in the previous section to construct an agent based stochastic version. In this model, the individual agents are assumed to be located on a two dimensional lattice of dimension300*×*300. The points on the lattice represent susceptible agents in the population and each individual on the lattice is surrounded by four nearest neighbors at a minimum distance of one. Here, we considered a radius of seven as the closest contacts of every agent on the lattice. Thus, every individual agent would be in contact with a maximum of 28 neighbors. The initial patient at zero time is located exactly at the center having a location of (150,150) on this two dimensional lattice. Infection spreads through persistent close contacts with the nearest neighbors. At every time step, all the exposed and the infected agents are selected and 28 neighbors of the individual get exposed to the infection with a probability *β*. The The value *β_ODE_*in the ODE model is defined as the number of individuals a particular person infects per unit time. In the stochastic model each individual would infect 28*×β* people on an average per unit time. Thus, the conversion between transmission rates of the ODE and stochastic model is given by 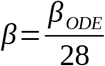. Here we assumed that both the asymptomatic/exposed and symptomatic patients are capable of spreading the infection. The exposed patient in turn exhibits symptom with a probability *α*_1_ and the symptomatic infected patient would be further quarantined with a probability *α*_2_. The quarantined patient finally either recovers or dies with a rate *γ*. The susceptible, exposed, infected, quarantined and recovered agents on the lattice are assigned values 0,1,2,3 and 4 respectively which allow us to track the dynamics of each component separately. The parameters values were taken from the fitted values of the ODE based model for a particular country to simulate the real scenario in that country. In order to introduce diagnostic testing, we identified all the symptomatic as well as the quarantined individuals from the pool at a particular time point and selected a fraction(*F_test_*) of the identified patients for contact tracing. The contacts of each selected individual were traced and diagnostic tests were conducted on them. The close contacts showing positive results were finally quarantined. This procedure is repeated for all the selected individuals. The status of the whole population is updated and the next time step is continued. The simulation was performed for 200 time points. In order to increase the number of testing, the value of *F_test_* were increased. However, we assumed that the testing procedure only commences after at least 100 individuals are already infected in the population in order to introduce an initial delay in responding to the situation. For every infected person, we also keep track of the number of individuals who are getting infected from that particular infected person until that person is quarantined. The number of people infected by the time defines the reproduction number for that individual. The *R*_0_value for the whole population is quantified by taking average over the reproduction numbers for all the persons in the population.

### Calculation of of *R*_0_ from the ODE based model

If we assume quarantine by testing following the agent based model described in the previous section, the exposed asymptomatic population would also be quarantined with a rate in addition to the symptomatic infected patients. The equations for this case would be

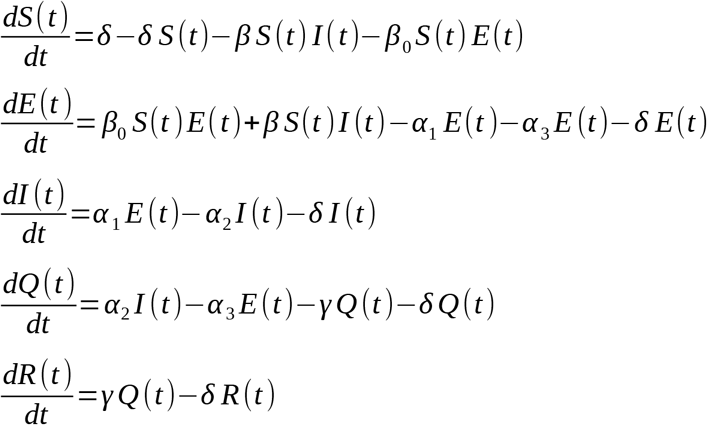

Where, we assumed a birth/death rate *δ* and an extra quarantine rate *α*_3_ in order to accommodate quarantine by testing which can also identify asymptomatic exposed individuals. In this calculation, we incorporated both the recovered and death by one equation for R(t).

The jacobian at the infection free equilibrium (I=0,E=0) is given by

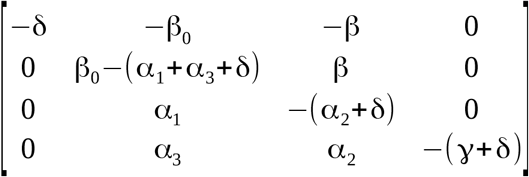

The eigenvalues are

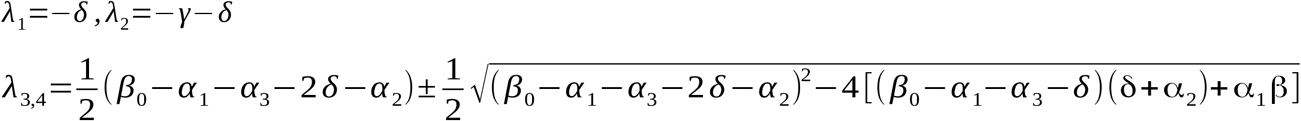

For the infection free equilibrium to be stable

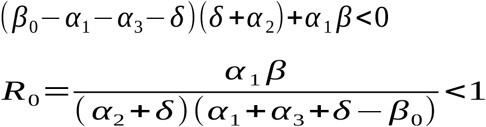

This shows that the reproduction ratio *R*_0_ would reduce if quarantine rate is increased by increasing the test rate through *α*_3_. In the original model for fitting, we assumed that *α*_3_ is small and additionally, the natural death rate/birth rate *δ*is also small in the time scale of the pandemic infection progression so that the total population characteristics remain the same over the timescale. The equation further demonstrates that *R*_0_ would also reduce if the incubation rate *α*_1_is high indicating the fact that if the asymptomatic exposed population exhibits symptoms at a higher rate, the spread of infection would be less. However, this will only happen when an asymptomatic carrier is capable of exposing a susceptible person. In fact, if *β*_0_is negligibly small (*β*_0_≪*α*_3_) in the equation above, the *R*_0_ value would no longer reduce with increasing *α*_1_. This result further illustrates that the effect of asymptomatic spread of infection can be largely mitigated by facilitating more testing and quarantine of the asymptomatic individuals.

#### A case for the periodic lockdown

In a previous study, it was suggested that a periodic lockdown may be a good strategy to optimize unlock and economic activity [3]. Thus, if after every *T*_1_time of full unlock, a lockdown period is imposed for a period of *T*_2_ over a total time N and this cycle continues. Hene, over a cycle of N, for the period *T*_1_, the 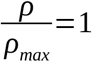 and followed by a period *T*_2_ where *ρ*=1. The average *R*_0_ over this period is given by

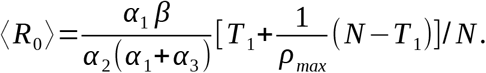

To keep the average equal to 1,

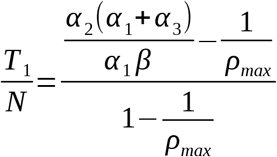

which represents allowed unlock number of days over a period time N to keep the average *R*_0_ at equal to 1. This shows that as the effect of lockdown *ρ* is high the allowed unlock period would also be high. The allowed period can further be stretched by increasing the quarantine rate involved with testing and quarantining the exposed subjects at a rate *α*_3_ where as small value of *α*_3_ corresponds to the case when testing rate is low which is used in fitting the data.

